# Impact of Public Health Education Program on the Novel Coronavirus Outbreak in the United States

**DOI:** 10.1101/2021.01.18.21250047

**Authors:** Enahoro A. Iboi, Ariana Richardson, Rachel Ruffin, DeAndrea Ingram, Jailyn Clark, Jala Hawkins, Maati McKinney, Nianza Horne, Reyla Ponder, Zoe Denton, Folashade B. Agusto, Bismark Oduro, Lanre Akinyemi

## Abstract

The coronavirus outbreak in the United States continues to pose a serious threat to human lives. Public health measures to slow down the spread of the virus involve using a face mask, social-distancing, and frequent hand washing. Since the beginning of the pandemic, there has been a global campaign on the use of non-pharmaceutical interventions (NPIs) to curtail the spread of the virus. However, the number of cases, mortality, and hospitalization continue to rise globally, including in the United States. We developed a mathematical model to assess the impact of a public health education program on the coronavirus outbreak in the US. Our simulation showed the prospect of an effective public health education program in reducing both the cumulative and daily mortality of the novel coronavirus. Finally, our result suggests the need to obey public health measures as loss of willingness would increase the cumulative and daily mortality in the US.

## 1 Introduction

The novel coronavirus (COVID-19) pandemic caused by SARS-CoV-2 was first reported in Wuhan, China in December 2019 and later declared a pandemic by the World Health Organization (WHO) on March 11, 2020 [1–3]. The emergence of the virus continues to cause devastating public health, and social-economic impact around the globe, including the United States (US) [4, 5]. COVID-19 is primarily transmitted from person-to-person through respiratory droplets released when an infected person sneezes, coughs, or talks [6]. The symptoms for COVID-19, which are similar to the common cold, though potentially more severe, include fever, cough, shortness of breath, fatigue, loss of taste or smell, sore throat, running nose, nausea, and diarrhea [6]. As of December 12, 2020, there are over 71 million confirmed COVID-19 cases globally, resulting in over 1.6 million deaths [7]. Within the United States, there have been over 16 million confirmed cases of coronavirus, with over 297,501 deaths [4].

The Centers for Disease Control and Prevention (CDC) on April 2, 2020, recommended the use of non-pharmaceutical interventions (NPIs) such as face masks in public (see Fig.1) and to practice social-distancing to curtail the spread of the virus [3, 5, 8–11]. Non-pharmaceutical interventions have had a long history of preventing many infectious diseases such as the pandemic Influenza, Measles, and the Ebola Virus Disease (EVD) [12–16]. Actions taken in the early stage of the coronavirus outbreak by the various state governments in the US include declaring a state of emergency and issuing a state-wide shelter in place. The use of a face mask by the general public in the US has been controversial as some state governors issued executive orders that voided face mask mandates within their jurisdiction [17].

Numerous mathematical models have been used to provide insights into public health measures for mitigating the spread of the novel coronavirus pandemic. Ferguson *et al*. [18] proposed an agent-based model to assess the impact of NPIs on COVID-19 mortality. In the absence of public health interventions, their model projected high mortality in the United States and the United Kingdom. Eikenberry *et al*. [3] developed a mathematical model to assess the impact of mask use by the general public on the transmission dynamics of the COVID-19 pandemic. Their results showed that broad adoption of even relatively ineffective face masks might reduce community transmission of COVID-19 and decrease peak hospitalizations and deaths. Recently, Ngonghala *et al*. [8] developed a mathematical model to assess the impact of NPIs on curtailing the public health burden of COVID-19 in the United States. Their study showed the effect of early implementation of face masks, lockdown, and lifting of social-distancing. Extending the duration of lockdown could reduce the daily cases, daily mortality in the US. Mizumoto and Chowell [19] used a mathematical model to assess the potential for a coronavirus outbreak aboard the Diamond Princess cruise, which experienced a major COVID-19 outbreak during the months of January and February of 2020. Their study showed that the basic reproduction number of the model decreases with increasing the effectiveness of the quarantine and isolation measures implemented on the ship.

Despite public health campaigns regarding the use of a face masks and social-distancing in the United States, the local transmission of COVID-19 throughout different parts of the country continues to rise. While many people follow public health recommendations to the use of face mask and practice social-distance in public to limit the spread of the virus, others passionately fight against them. It is important to understand how educating the population on the importance of using a face mask and social-distancing could reduce the spread of the virus. The objective of this study is to use a mathematical model to assess the impact of public health education campaigns on the coronavirus outbreak in the United States.

## 2 Materials and Methods

### 2.1 Model Formulation

The coronavirus model to be developed uses the natural history of the infection. The total human population at time *t*, denoted by *N* (*t*), is sub-divided into mutually exclusive compartments of unwilling susceptible (*S*_*u*_(*t*)), willing susceptible (*S*_*e*_(*t*)), unwilling exposed (*E*_*u*_(*t*)), willing exposed (*E*_*e*_(*t*)), unwilling asymptomatic-infectious (*A*_*u*_(*t*)), willing asymptomatic-infectious (*A*_*e*_(*t*)), unwilling infectious with symptoms (*I*_*us*_(*t*)), willing infectious with symptoms (*I*_*es*_(*t*)), unwilling hospitalized or isolated at a health care facility (*H*_*u*_(*t*)), willing hospitalized or isolated at a health care facility (*H*_*e*_(*t*)), in intensive care units (*I*_*cu*_(*t*)), and recovered (*R*(*t*)) individuals. Thus, the total population size *N* is given as

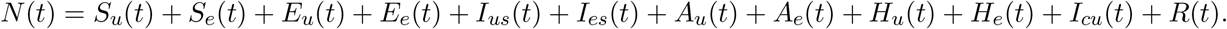

The flow diagram of the model (2.1) is depicted in Figure 2 (the state variables and parameters of the model are described in Tables 1 and 2, respectively).

**Table 1:** Description of the state variables of the model (2.1)

| State variable | Description |
| --- | --- |
| $S_u$ | Population of unwilling susceptible individuals |
| $S_e$ | Population of willing susceptible individuals |
| $E_u$ | Population of unwilling exposed individuals |
| $E_e$ | Population of willing exposed individuals |
| $I_{us}$ | Population of unwilling infectious individuals with severe clinical symptoms of COVID-19 |
| $I_{es}$ | Population of willing infectious individuals with severe clinical symptoms of COVID-19 |
| $A_u$ | Population of unwilling asymptomatic-Infectious individuals |
| $A_e$ | Population of willing asymptomatic-Infectious individuals |
| $H_u$ | Population of unwilling hospitalized individuals |
| $H_e$ | Population of willing hospitalized individuals |
| $I_{cu}$ | Population of individuals in ICU |
| $R$ | Population of recovered individuals |

**Table 2:** Description of parameters of the model (2.1)

| Parameter | Description |
| --- | --- |
| $\beta$ | Effective contact rates for willing(unwilling) individuals |
| $\omega$ | Efficacy of education in preventing COVID-19 infection ($0 < \omega \leq 1$) |
| $\eta_{A_u}(\eta_{A_e})(\eta_{H_u})(\eta_{H_e})$ | Modification parameters ($0 < \eta_{A_u}(\eta_{A_e})(\eta_{H_u})(\eta_{H_e}) < 1$) |
| $\psi$ | Education rate for individuals in $S_u$ ($E_u$) ($I_{us}$) ($A_u$) |
| $\nu$ | Fatigue rate (loss of willingness to public health measures) |
| $\sigma_u(\sigma_e)$ | Progression rates from $E_u$ ($E_e$) to $I_{us}$ ($I_{es}$) or $A_e$ ($A_e$) class |
| $r(g)$ | Proportion of individuals in $E_u$ ($E_e$) class who show clinical symptoms of COVID-19 |
| $\alpha_{us}(\alpha_{es})$ | Hospitalization rates for unwilling(willing) infectious individuals |
| $\phi_{hu}(\phi_{he})$ | ICU admission rate for unwilling(willing) hospitalized individuals |
| $\gamma_{ua}(\gamma_{ea})(\gamma_{us})(\gamma_{es})(\gamma_h)(\gamma_{cu})$ | Recovery rates for individuals in the $A(I_s)(H)(I_{cu})$ class |
| $\delta_{us}(\delta_{es})(\delta_h)(\delta_{cu})$ | Disease-induced death rates for individuals in the $I_{us}(I_{es})(H)(I_{cu})$ class |

**Figure 1:**
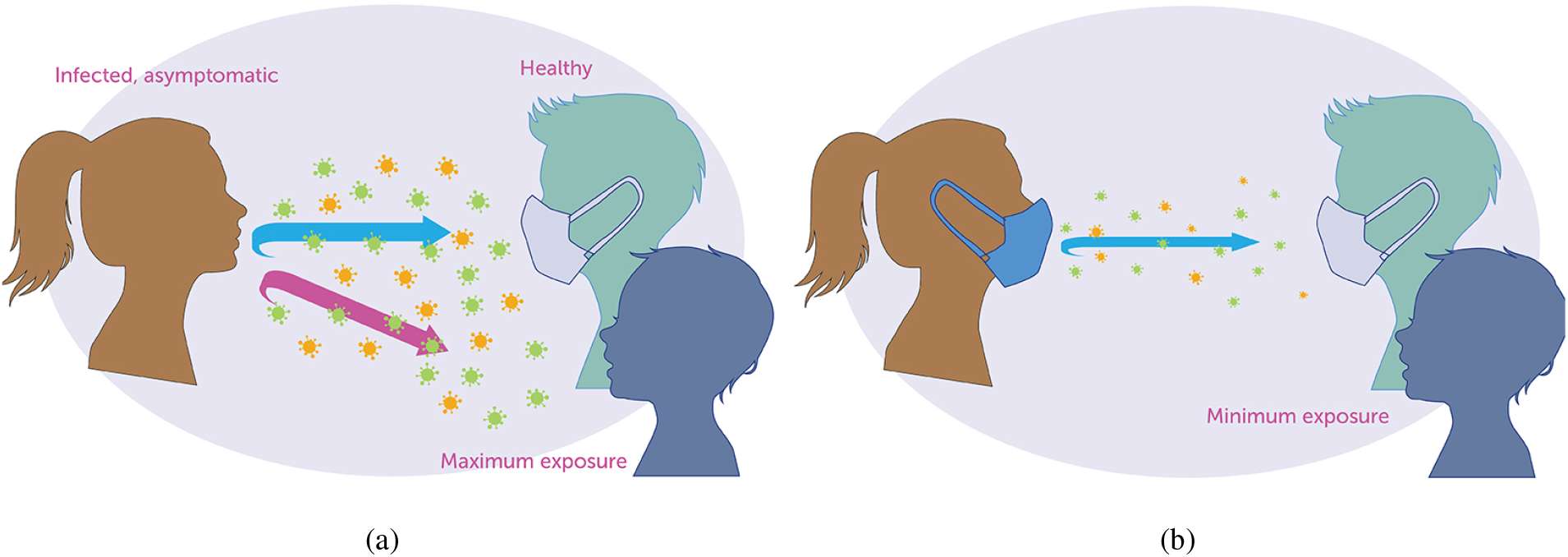
The science of mask against COVID-19 [20].

**Figure 2:**
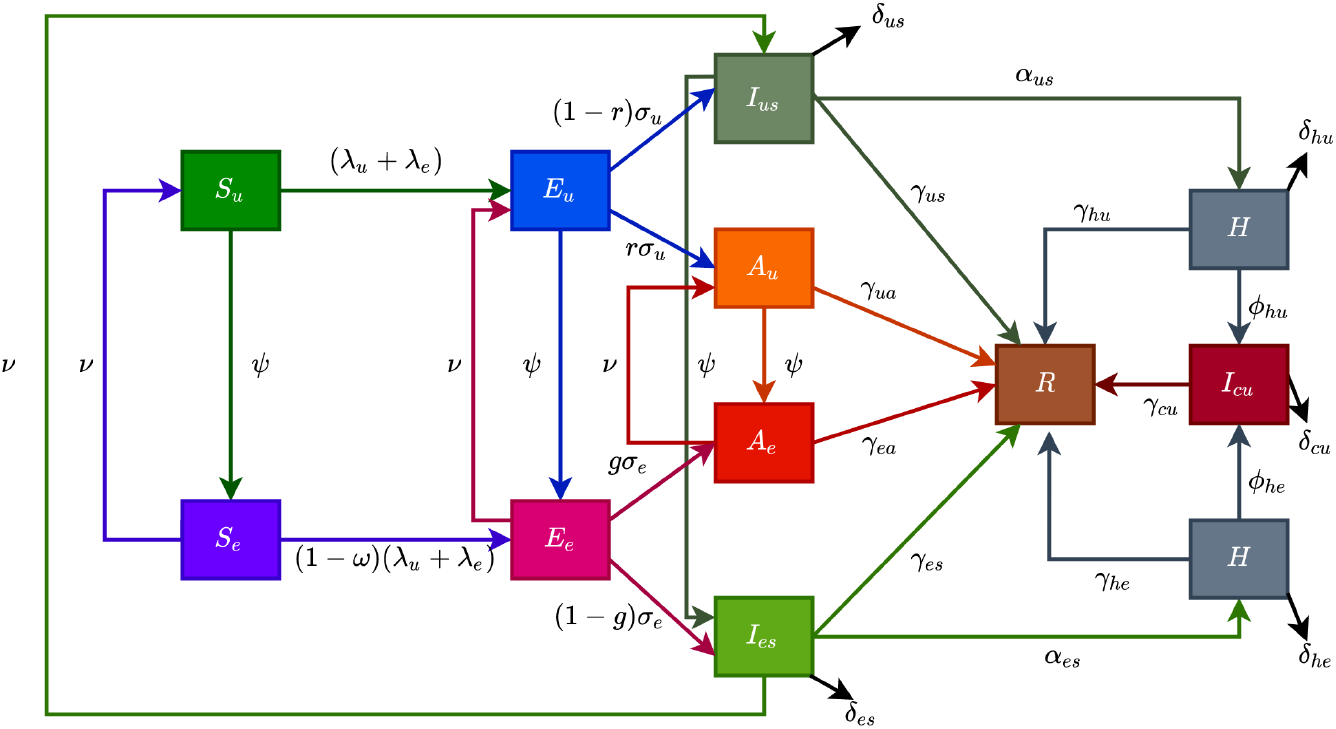
Flow diagram of the model (2.1) showing transitions from various compartments based on public health education.

The model is given by the following deterministic system of nonlinear differential equations (where a dot represents differentiation with respect to time *t*):

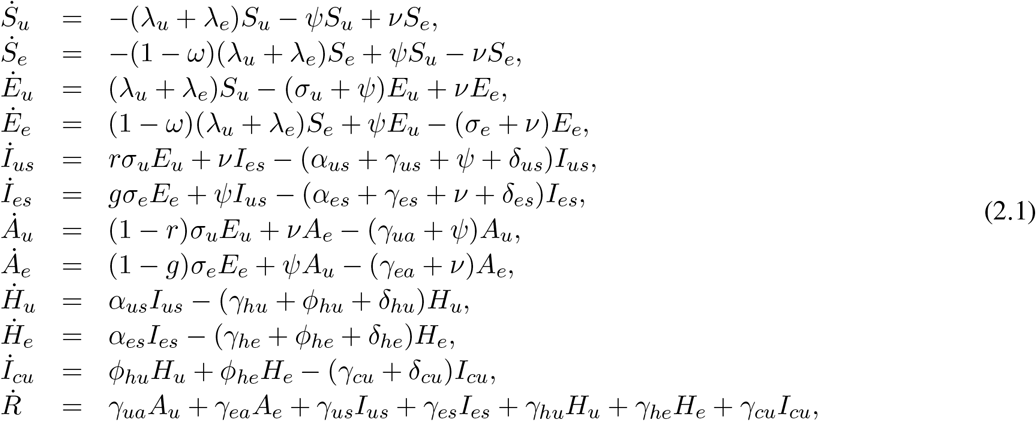

where the forces of infection are given by

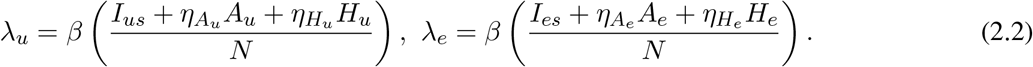

In model 2.1, *β* is the effective infection rate for unwilling and willing individuals, while *η*_*j*_, (*j* ∈ {*A*_*k*_, *H*_*k*_}, *k* ∈ {*u, e*}), is the modification parameters (where 0 *< η*_*j*_ *<* 1) that accounts for a reduction in infectiousness of unwilling(willing) asymptomatic and hospitalized individuals compared to unwilling(willing) symptomatic individuals. Further, *ψ* represent the public health education rate for unwilling susceptible (*S*_*u*_), exposed (*E*_*u*_), and symptomatic individuals (*I*_*us*_), and asymptomatic (*A*_*u*_) respectively. It is assumed that public health education program towards the use of NPIs in preventing COVID-19 infection is imperfect (i.e., allowing willing susceptible individuals become infected with COVID-19), with an efficacy *ω* (where 0 *< ω* ≤ 1). Furthermore, the parameters *σ*_*j*_, *j* = *u, e* represents the progression rates of unwilling (willing) exposed individuals. A proportion, 0 *< r, g* ≤ 1, of unwilling (willing) exposed individuals show clinical symptoms of COVID-19 and move to the class *I*_*js*_, *j* = *u, e*, at the end of the incubation period. The remaining proportion, (1 − *r*) and (1 − *g*), show no clinical symptoms and move to the *A*_*j*_, *j* = *u, e*, class. Further, *ν* represent the loss of willingness to wear a face mask, practice social-distancing in public, and frequently washing hands. The parameters *α*_*js*_, *j* = *u, e*, is the hospitalization (or self-isolation) rates of unwilling(willing) individuals with clinical symptoms of COVID-19. Similarly, the parameters *ϕ*_*hu*_, *ϕ*_*he*_ is the ICU admission rates. The parameters *γ*_*ja*_, *γ*_*js*_, *γ*_*hj*_, *γ*_*cu*_, *j* = *u, e*, represents the recovery rates for unwilling (willing) individuals in the *A*_*j*_, *I*_*js*_, *H*_*j*_, *I*_*cu*_, *j* = *u, e* classes. Finally, the parameter *δ*_*js*_, *δ*_*hj*_, *δ*_*cu*_, *j* = *u, e* represents the COVID-induced mortality rate for individuals in the *I*_*js*_, *H*_*j*_, *I*_*cu*_, *j* = *u, e* classes. To formulate the model, we made the following assumptions:

i. due to public health education, willing individuals wear face mask to prevent transmission, practise socialdistancing and wash their hands while unwilling individuals do not.
ii. public health education program is targeted at individuals who are unwilling to use a face mask or practice social-distance in public at rate (*ψ*).
iii. to account for public health education saturation, we assume a willingness fatigue (i.e., loss of willingness to wear face mask, practise social-distancing, and frequent washing of hands),

The model (2.1) is also an extension of the COVID-19 models in [3, 5, 8–10] by including compartments for individuals based on their willingness/unwillingness regarding the adherence to non-pharmaceutical interventions such as face mask, social-distancing, and hand washing to curtail the COVID-19 outbreak. Models of this type have been formulated for Influenza [12].

## 3 Results

### 3.1 Asymptotic Stability of Disease-free Equilibria

The expression for the reproduction number (ℛ_*c*_) for model with public health education program is given in Appendix B.

#### Theorem 3.1.

*The disease-free equilibrium (DFE) of the model* (2.1) *is locally-asymptotically stable if* ℛ_*c*_ *<* 1. *If* ℛ_*c*_ *>* 1, *the epidemic grows rapidly, reaches a peak, and eventually declines to zero*.

The quantity ℛ_*c*_ is the *reproduction number* of the model (2.1). It measures the average number of new COVID-19 cases generated by a typical infectious individual introduced into a population where a certain fraction is protected.

### 3.2 Data Fitting and Parameter Estimation

Estimates for some of the parameters of the model (2.1) were obtained from the literature (as indicated in Table 3). Other parameters, such as the effective infection rate parameters *β*, education rate *ψ*, education efficacy *ω*, and fatigue rate *ν* are obtained by fitting the model to the observed cumulative mortality data for the US [21, 22]. In particular, the US Cumulative mortality data from January 22, 2020 (first index case) to December 8, 2020 were obtained from the John Hopkins’ Center for Systems Science and Engineering COVID-19 Dashboard [23]. We fitted the model for three different time periods of the pandemic, with the first period from January 22, 2020 to July 5, 2020, second period from July 6, 2020 to September 30, 2020 and the third period from October 1, 2020 to December 8, 2020. This was done in order to correctly capture the trends observed in the daily mortality data (i.e., the COVID-19 waves observed). Hence, we obtained three set of values for the parameters to be estimated based on the different periods. Our choice of fitting the model to the mortality data is due to the fact that there is evidence of under-reporting and under-testing of COVID-19 cases in countries such as France, Italy, United States, Iran, and Spain. Hence, mortality data may provide a better indicator for COVID-19 case spread [8, 24]. The data-fitting process involves implementing the standard nonlinear least squares approach using the *fmincon* Optimization Toolbox embedded in MATLAB. The estimated values of the unknown parameters are tabulated in Table 4. Figure 3(a)-3(c) depicts the fitting of the observed and predicted cumulative mortality for the US. Further, Figure 3(d)-3(e) compares the simulations of the model using the fitted and fixed parameter in Tables 3 and 4. The results depicted in Fig. 3, show that the model also captures the observed daily mortality data for each of the period considered. Thus, the parameter estimation of model (2.1) shows that cumulative mortality data provides a very reliable calibration for coronavirus transmission dynamics.

**Table 3:** Baseline parameter values for the model (2.1) drawn from the literature.

| Fixed Parameter ($k = u, e$) | Value | Reference |
| --- | --- | --- |
| $\sigma_e, \sigma_u$ | 1/2.5/day | [31, 32] |
| $r, g$ | 0.35 | [33, 34] |
| $\eta_{A_k}$ | 1.5 | Assumed |
| $\eta_{H_k}$ | 0.25 | Assumed |
| $\alpha_{us}, \alpha_{es}$ | 1/6/day | [35] |
| $\phi_{hu}, \phi_{he}$ | 0.083/day | [36] |
| $\gamma_{ua}, \gamma_{ea}$ | 1/5/day | [35] |
| $\gamma_{us}, \gamma_{es}$ | 1/10/day | [18, 37] |
| $\gamma_{hu}, \gamma_{he}$ | 1/8/day | [18] |
| $\gamma_{cu}$ | 1/10/day | [18, 37] |
| $\delta_{ks}$ | 0.015/day | [3, 5, 18] |
| $\delta_{hk}$ | 0.015/day | [3, 5, 18] |
| $\delta_{cu}$ | 0.0225/day | [3, 5, 18] |

**Table 4:** Estimated parameter values for the model (2.1) using COVID-19 mortality data for the US.

| Estimated Parameters | 1/22/2020–7/5/2020 | 7/6/2020–9/30/2020 | 10/1/2020–12/8/2020 |
| --- | --- | --- | --- |
| $\beta$ | 0.8084 | 0.4369 | 0.2842 |
| $\psi$ | 0.0279 | 0.0781 | 0.0249 |
| $\nu$ | 0.0011 | 0.0210 | 0.0461 |
| $\omega$ | 0.8982 | 0.8599 | 0.8896 |

**Figure 3:**
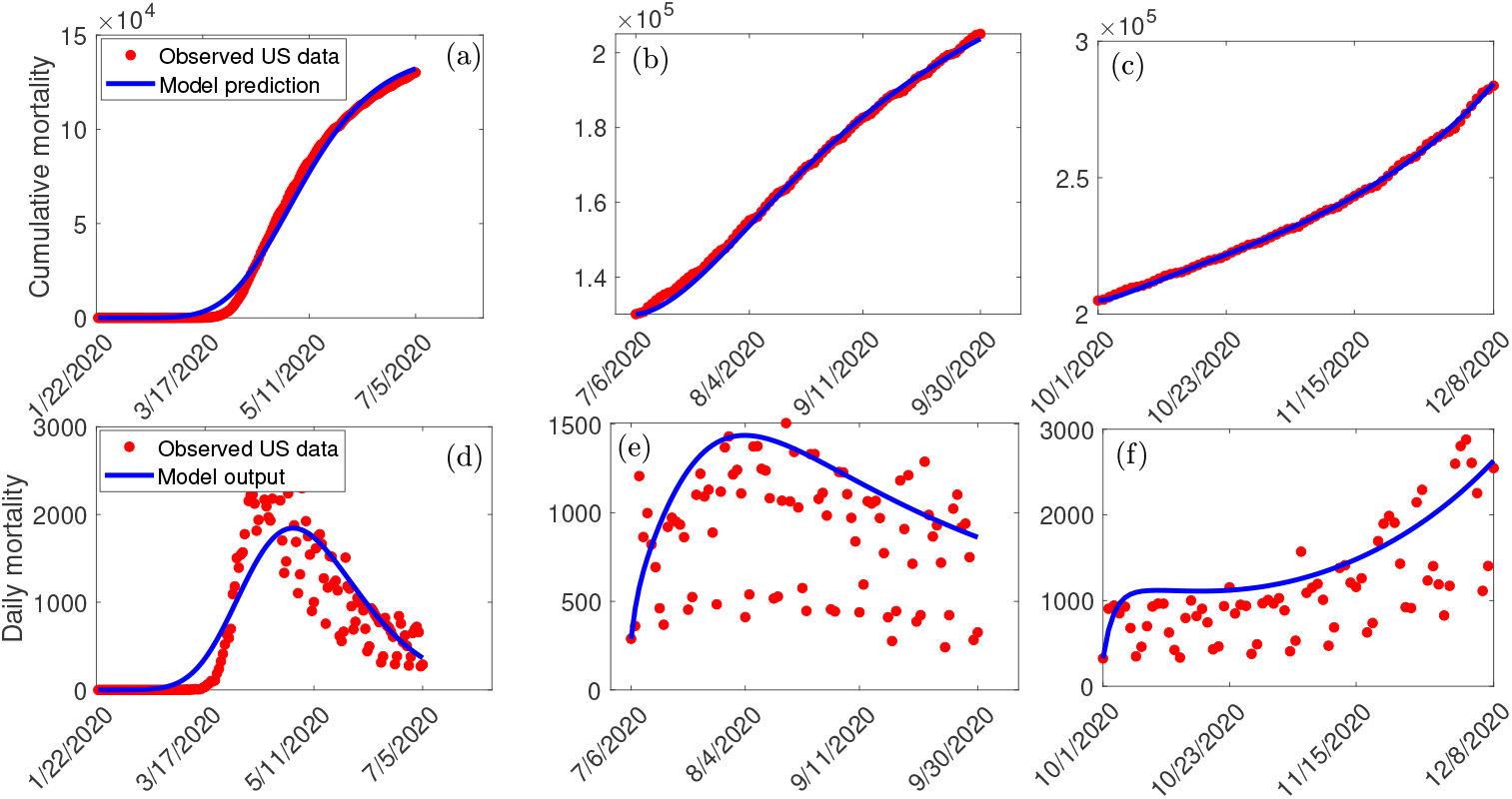
(a)-(c): Data fitting of the model (2.1) using the cumulative mortality data for the US from January 22, 2020 to December 8, 2020. (d)-(f): Simulations of the model (2.1) using the fixed and the fitted parameters from the cumulative mortality data for the US in Tables 3 and 4.

### 3.3 Sensitivity Analysis

The model (2.1) contains 26 parameters, and uncertainty in their estimates are expected to arise. The effect of such uncertainties is assessed using uncertainty and sensitivity analysis [25–27]. In particular Latin Hypercube Sampling (LHS) and Partial Rank Correlation Coefficients (PRCC) is used to identify model parameters that have the most influence on the model with the reproduction number (ℛ_*c*_) as the response function. The purpose of this analysis is to determine effects of parameters on model outcomes [25–27]. A highly sensitive parameter should be more carefully estimated, since a small change in that parameter can cause a large quantitative changes in the result [25–27]. On the other hand, a parameter that is not sensitive does not require as much attempt to estimate, since a small change in that parameter will not cause a large variation to the quantity of interest [26]. Parameters with large PRCC greater than +0.50 are said to be highly positively correlated with the response function, while those less than −0.50 are said to be highly negatively correlated with the response function [25–27]. The parameters considered in the PRCCs analysis are the effective infection rate for unwilling(willing) individuals (*β*), education rates for unwilling(willing) individuals (*ψ*), education efficacy (*ω*), and fatigue rate (*ν*). We performed a PRCC analysis for the three different periods; however, the parameters have the same effect on the response function for the three periods. We chose to report one plot as displayed in Figure 4. The results show that the parameters that mostly impact the response function (ℛ_*c*_) are the effective infection rate (*β*), education rate (*ψ*), fatigue rate (*ν*), and education efficacy (*ω*). Based on the PRCC values, the transmission rate for unwilling individuals and the fatigue rate has a positive impact on ℛ_*c*_, as an increase(decrease) in the transmission and fatigue parameter will increase(decrease) ℛ_*c*_. In contrast, the education rate and efficacy have a negative impact on the ℛ_*c*_, and an increase in these parameters will decrease the ℛ_*c*_.

**Figure 4:**
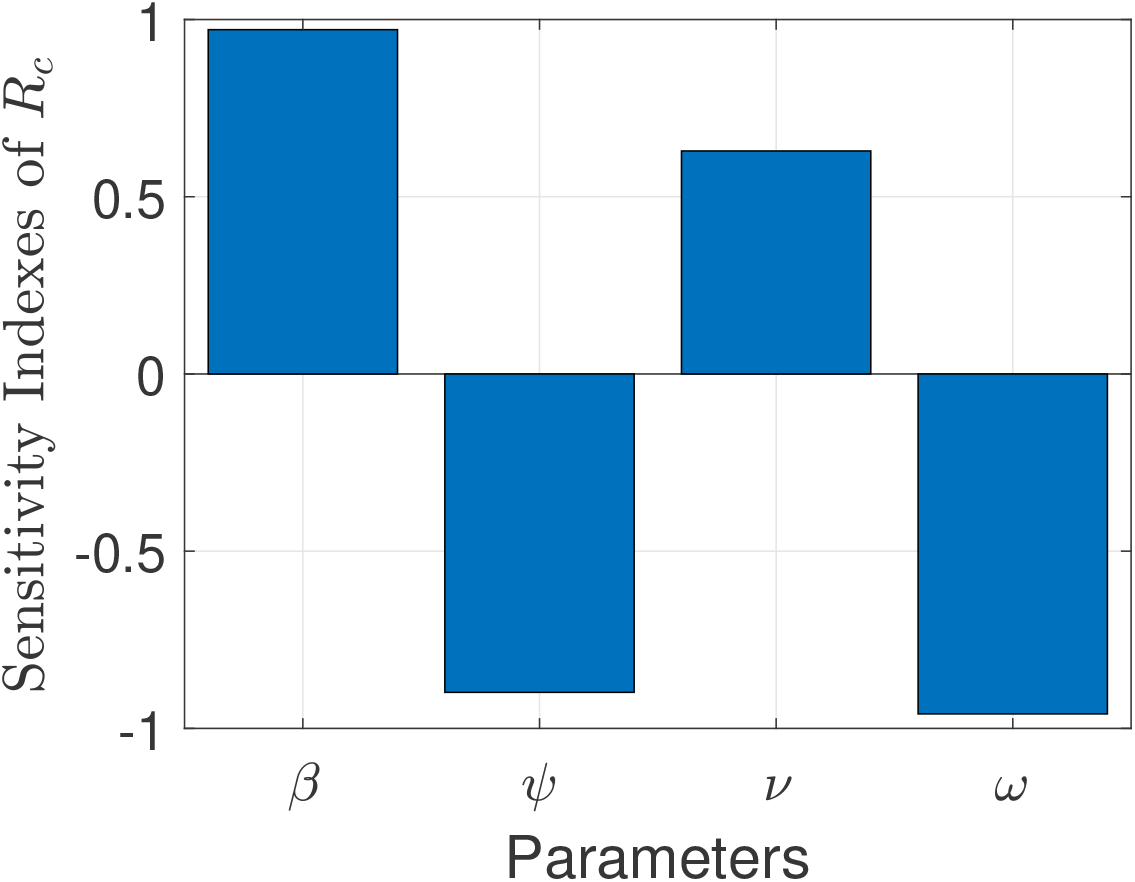
Partial rank correlation coefficients (PRCCs) showing the impact of model parameters on the reproduction number (ℛ_*c*_) of the model. Parameter values used are as given in Tables 3 and 4.

### 3.4 Numerical Simulation Results

To capture the trends observed in the daily mortality data obtained for the US from January 22, 2020, to December 8, 2020, we considered three different periods of the pandemic with the first period from January 22, 2020, to July 5, 2020, second period from July 6, 2020 to September 30, 2020, and the third period from October 1, 2020 to December 8, 2020. First, we generated a contour plot of the reproduction number (ℛ_*c*_) of the model (2.1), as a function of education rate (*ψ*) and education efficacy (*ω*) (Fig. 5). Figure 5(a) shows that for the period January 22, 2020 to July 5, 2020, a very high education rate (*ψ*) is necessary to curtail the outbreak effectively (i.e., bring ℛ_*c*_ to a value less than one). A similar trend is observed for the period July 6, 2020, to September 30, 2020, of the outbreak (Fig. 5(b)). However, Fig. 5(c) shows that for the period October 1, 2020, to December 8, 2020, as more people are being educated with high efficacy, the value of ℛ_*c*_ decreases. It is worth mentioning that the value of ℛ_*c*_ depends on the initial conditions, more precisely on the location of the specific DFE within the hyperplane of disease-free equilibria. Assuming that no individuals are educated at the beginning of the simulation, then the education efficacy (*ω*) will be irrelevant (sensitivity index close to zero) since there are no individuals being educated. As soon as a significant number of individuals is educated, the effect of the education efficacy on ℛ_*c*_ will increase dramatically. Moreover, since ℛ_*c*_ depends on the values of the initial conditions (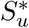 and 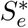), it is expected that ℛ_*c*_ decreases as more individuals are being educated over time.

**Figure 5:**
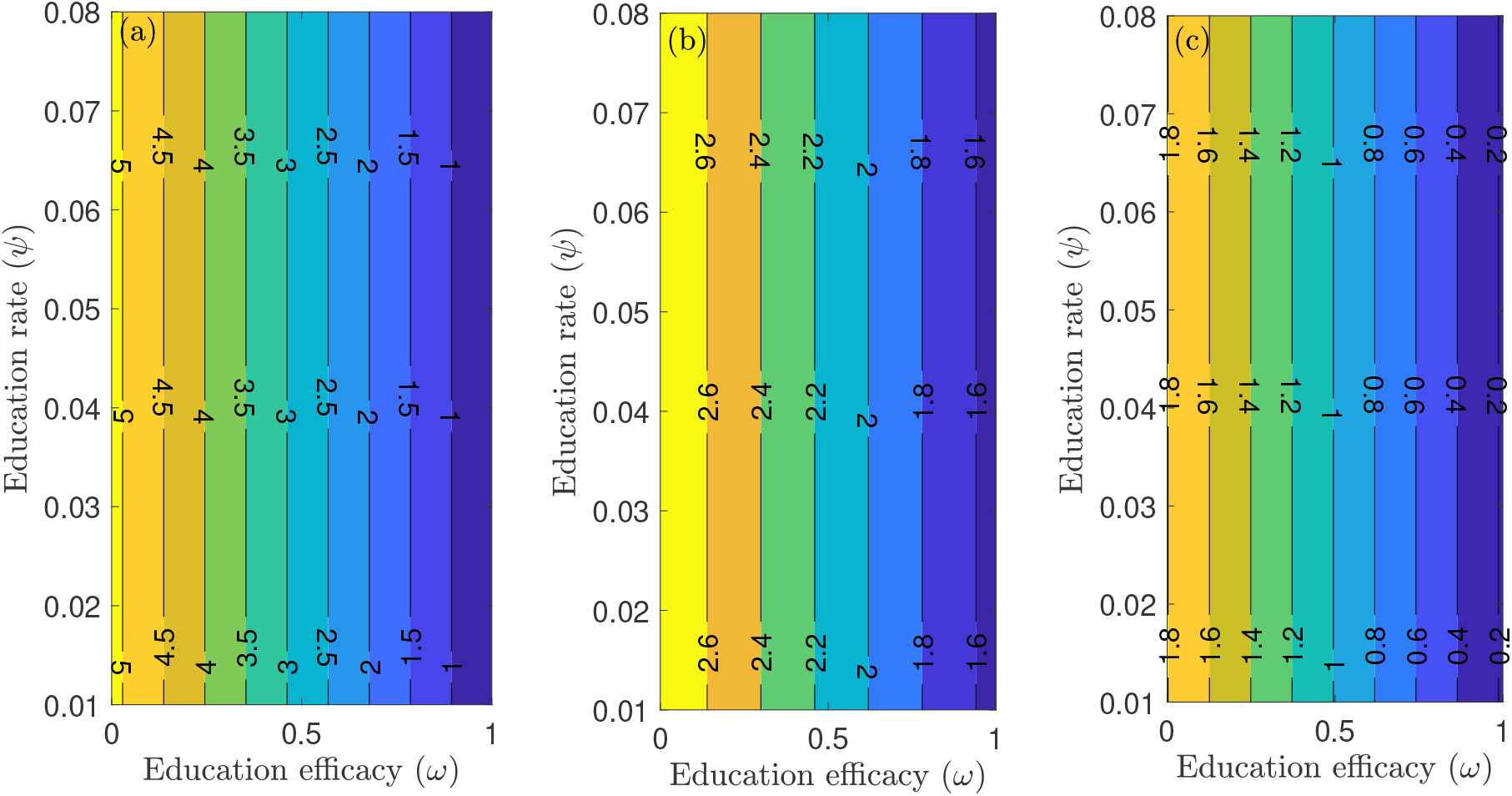
Contour plot of the reproduction number (ℛ_*c*_) of the model (2.1), as a function of education rate (*ψ*) and education efficacy (*ω*). (a) First period from January 22, 2020 to July 5, 2020. (b) Second period from July 6, 2020 to September 30, 2020. (c) Third period from October 1, 2020 to December 8, 2020. Parameter values are as given in Tables 3 and 4.

Figure 6 depicts a contour plot of the reproduction number (ℛ_*c*_) of the model (2.1), as a function of the proportion of educated individuals among all susceptible 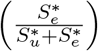 and education efficacy (*ω*) for a fixed education rate (*ψ*). Figure 6(a) shows that for the period January 22, 2020 to July 5, 2020, with the baseline education efficacy, ℛ_*c*_ can be brought to a value less than one if 90% among all susceptible individuals are educated. This result suggests that an incredibly high education rate (*ψ*) is necessary to curtail the outbreak effectively for the period January 22, 2020, to July 5, 2020. However, for the period July 6, 2020, to September 30, 2020, of the outbreak, with the baseline education efficacy, ℛ_*c*_ can be brought to a value less than one if 76% among all susceptible individuals are educated (Fig. 6(b)). Figure 6(c) shows that for the period October 1, 2020, to December 8, 2020, with the baseline education efficacy, ℛ_*c*_ can be brought to a value less than one if 51% among all susceptible individuals are educated. This result further supports the need to educate more people if we are to effectively curtail the coronavirus outbreak, which is consistent with the results obtained in Fig. 5.

**Figure 6:**
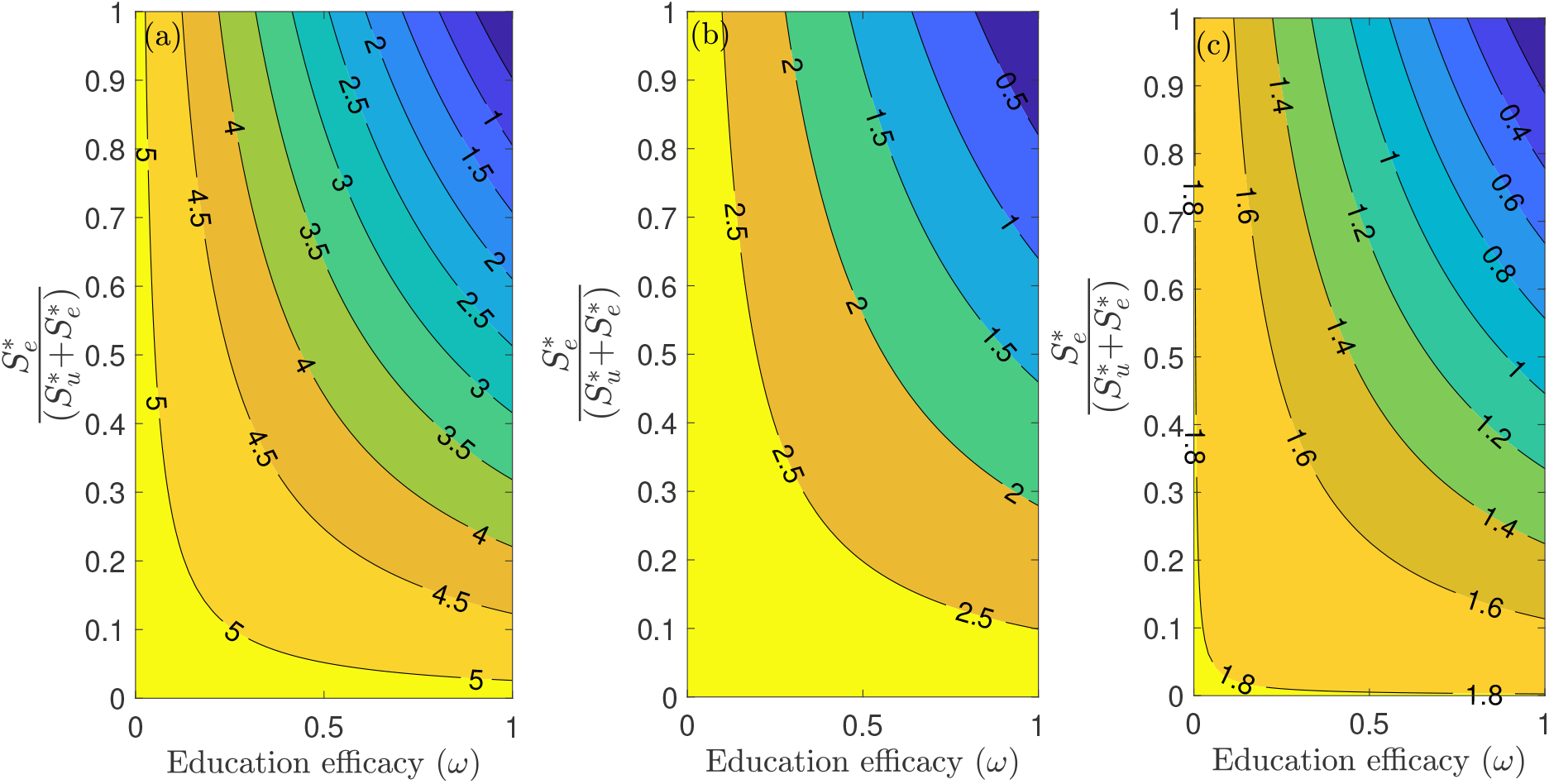
Contour plot of the reproduction mumber (ℛ_*c*_) of the model (2.1), as a function of the proportion of educated individuals among all susceptible 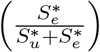 and education efficacy (*ω*) with a fixed education rate *ψ*. (a) First period from January 22, 2020 to July 5, 2020. (b) Second period from July 6, 2020 to September 30, 2020. (c) Third period from October 1, 2020 to December 8, 2020. Parameter values are as given in Tables 3 and 4.

Furthermore, we ran simulations of model (2.1) using the parameter values in Tables 3 and 4, to assess the population-level impact of public health education program on the COVID-19 outbreak. The simulation result for the baseline scenario shows a projected 132,000 cumulative deaths by July 5, 2020, 205,600 by September 30, 2020, and 285,100 by December 8, 2020 (Fig. 7(a)). Similarly, the projected peak daily mortality was 1,829 attained on April 28, 2020, 1,505 attained on August 3, 2020, and 2,808 attained by December 8, 2020 (Fig. 7(b)). Further, with a 10% increase in education rate from the baseline value, Fig. 7(a) shows a projected 44,400 cumulative mortality by July 5, 2020, 105,300 by September 30, 2020, and 178,000 by December 8, 2020. This result is approximately a 66.4% reduction in cumulative mortality by July 5, 2020, a 48.8% reduction in cumulative mortality by September 30, 2020, a 37.6% reduction in cumulative mortality by December 8, 2020, when compared to the baseline scenario. Figure 7(b) with a 10% increase in education rate from the baseline value, shows projected 617 peak mortality by April 20, 2020, 1,103 by July 31, 2020, and 2,359 by December 8, 2020. This result is approximately a 66.3% reduction in peak daily mortality by April, 20, 2020, a 26.7% reduction in peak daily mortality by July 31, 2020, a 16% reduction in peak daily mortality by December 8, 2020 when compared to the baseline scenario. However, with a 40% increase in education rate from the baseline value, Fig. 7(a) shows a projected 3,835 cumulative mortality by July 5, 2020, 44,410 by September 30, 2020, and 102,200 by December 8, 2020. This result is approximately a 97.1% reduction in cumulative mortality by July 5, 2020, a 78.4% reduction in cumulative mortality by September 30, 2020, a 64.2% reduction in cumulative mortality by December 8, 2020, when compared to the baseline scenario. Figure 7(b) with a 40% increase in education rate from the baseline value, shows projected 58 peak mortality by April 7, 2020, 836 by July 24, 2020, and 1,268 by December 8, 2020. This result is approximately a 96.8% reduction in peak daily mortality by April, 7, 2020, a 44.5% reduction in peak daily mortality by July 24, 2020, a 54.8% reduction in peak daily mortality by December 8, 2020, when compared to the baseline scenario. The result in Fig. 7 shows the need for an aggressive public health education program towards the use of NPIs to curtail the spread of the virus. A summary of the impact of various increase in education rate on cumulative mortality and peak daily mortality is tabulated in Table 5.

**Table 5:** A summary of various increase in education rate

| Education rate | 1/22/2020–7/5/2020 |  | 7/6/2020–8/30/2020 |  | 10/1/2020–12/8/2020 |  |
| --- | --- | --- | --- | --- | --- | --- |
|  | cum. mort. | daily mort. | cum. mort. | daily mort. | cum. mort. | daily mort. |
| Baseline | 132,000 | 1,829 | 205,600 | 1,505 | 285,100 | 2,808 |
| 10% increase in $\psi$ | 44,400 | 617 | 105,300 | 1,103 | 178,000 | 2,359 |
| 20% increase in $\psi$ | 17,210 | 248 | 69,160 | 962 | 136,200 | 1,867 |
| 30% increase in $\psi$ | 7,676 | 115 | 53,060 | 887 | 115,200 | 1,526 |
| 40% increase in $\psi$ | 3,835 | 58 | 44,410 | 836 | 102,200 | 1,268 |

**Figure 7:**
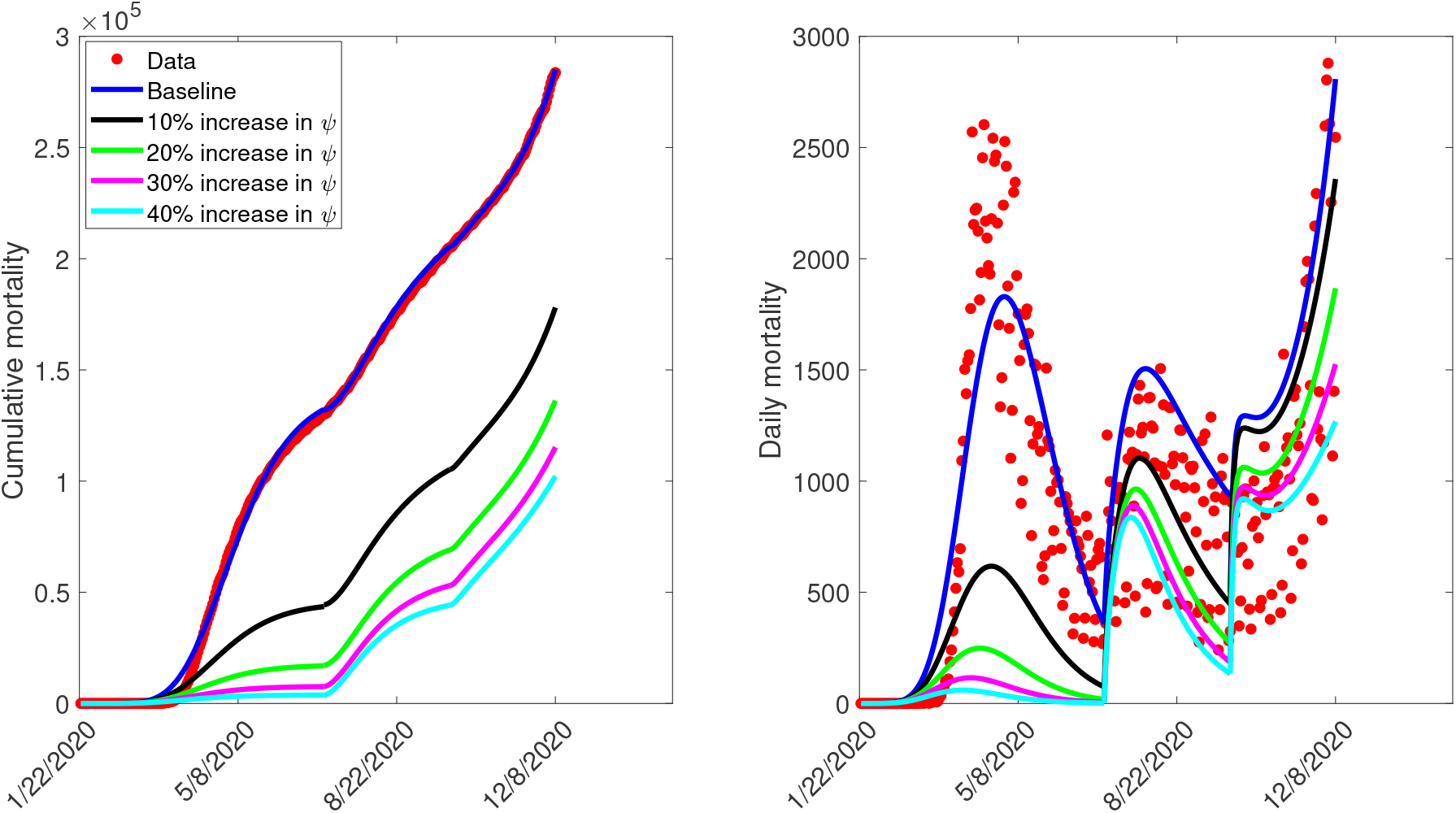
Simulation of the model (2.1), showing (a) cumulative mortality (b) daily deaths, as a function of time for various public health education rates in the US from January 22, 2020 to December 8, 2020. Parameter values are as given in Table 3.

Figure 8 depicts the impact of the loss of willingness to public health measures on COVID-19 outbreak. The result shows that with a 10% increase in fatigue rate from the baseline value, Figure 8(a) projected 144,100 cumulative mortality by July 5, 2020, 228,800 by September 30, 2020, and 325,700 by December 8, 2020. This result is approximately a 9.2% increase in cumulative deaths by July 5, 2020, a 11.3% increase in cumulative deaths by September 30, 2020, and a 14.2% increase in cumulative deaths by December 8, 2020, when compared to the baseline scenario. Figure 8(b) with a 10% increase in fatigue rate from the baseline value, shows projected 1,955 peak mortality by May 1, 2020, 1,657 by August 3, 2020, and 4,451 by December 8, 2020. This result is approximately a 6.9% increase in peak daily mortality by May 1, 2020, a 10.1% increase in peak daily mortality by August 3, 2020, a 58.5% increase in peak daily mortality by December 8, 2020, when compared to the baseline scenario. However, with a 40% increase in fatigue rate from the baseline value, Figure 8(a) shows a projected 184,200 cumulative mortality by July 5, 2020, 313,200 by September 30, 2020, and 478,300 by December 8, 2020. This result is approximately a 39.5% increase in cumulative deaths by July 5, 2020, a 52.3% increase in cumulative deaths by September 30, 2020, and a 67.8% increase in cumulative deaths by December 8, 2020 when compared to the baseline scenario. Figure 8(b) with a 40% increase in fatigue rate from the baseline value, shows projected 2,412 peak mortality by May 3, 2020, 2,513 by August 23, 2020, and 9,935 by December 8, 2020. This result is approximately a 31.9% increase in peak daily mortality by April, 20, 2020, a 67% increase in peak daily mortality by July 27, 2020, a 254% increase in peak daily mortality by December 8, 2020, when compared to the baseline scenario. This result suggests the need to obey public health measures as “loss of willingness” would positively impact the outcome of the pandemic in terms of the cumulative and the daily mortality in the US. A summary of the impact of the various increase in fatigue rate on cumulative mortality and peak daily mortality is tabulated in Table 6.

**Table 6:** A summary of various increase in fatigue rate

| Fatigue rate | 1/22/2020–7/5/2020 |  | 7/6/2020–8/30/2020 |  | 10/1/2020–12/8/2020 |  |
| --- | --- | --- | --- | --- | --- | --- |
|  | cum. mort. | daily mort. | cum. mort. | daily mort. | cum. mort. | daily mort. |
| Baseline | 132,000 | 1,829 | 205,600 | 1,505 | 285,100 | 2,808 |
| 10% increase in $\nu$ | 144,100 | 1,955 | 228,800 | 1,657 | 325,700 | 4,451 |
| 20% increase in $\nu$ | 157,400 | 2,094 | 253,200 | 1,860 | 370,300 | 5,953 |
| 30% increase in $\nu$ | 169,100 | 2,247 | 280,600 | 2,139 | 420,400 | 7,771 |
| 40% increase in $\nu$ | 184,200 | 2,412 | 313,200 | 2,513 | 478,300 | 9,935 |

**Figure 8:**
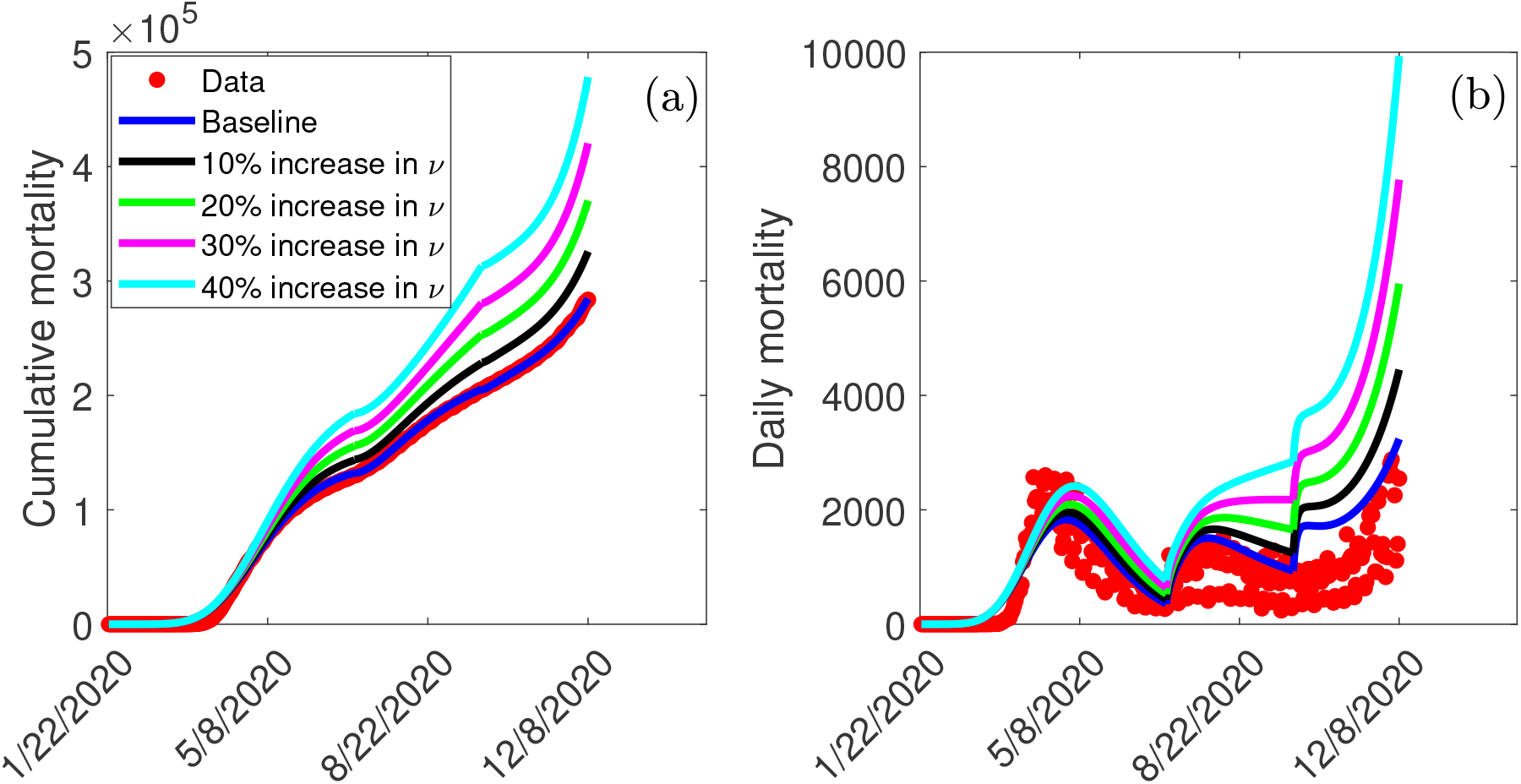
Simulation of the model (2.1), showing (a) cumulative mortality (b) daily deaths, as a function of time for various fatigue rates in the US from January 22, 2020 to December 8, 2020. Parameter values are as given in Table 3.

## 4 Discussion and Conclusions

In this study, we developed a mathematical model for the transmission dynamics and control of COVID-19 in the US by stratifying the total population into two subgroups of willing and unwilling individuals to the use of face masks, social-distancing in public, and proper/frequent hand washing. The model allows for the assessment of the impact of public health education programs on the coronavirus outbreak in the US. The model was parameterized using cumulative mortality data for the US from January 22, 2020, to December 8, 2020, to assess the population-level impact of public health education programs on the outbreak. In particular, we showed that the disease-free equilibrium of the model is locally-asymptotically stable whenever a certain epidemiological threshold, known as the reproduction number (ℛ_*c*_) is less than one. The epidemiological implication of this result is that when ℛ_*c*_ *<* 1, a small COVID-infected individuals in the community will not lead to an outbreak.

We explored the sensitivity of the reproduction number with respect to public health education rate in the US for three different periods of the outbreak. In particular, we showed that community transmission of COVID-19 could be significantly reduced with a very high education rate. In other words, our study shows that COVID-19 could have been effectively controlled if the public health education campaign has been intensified enough with high efficacy (and sustained) from the beginning of the pandemic. Furthermore, we also explored the sensitivity of the reproduction number with respect to willingness fatigue rate in the US for three different periods of the outbreak. Since the reproduction number ℛ_*c*_ depends on the values of the initial conditions (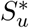 and 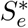), our result shows that ℛ_*c*_ can be brought to a value less than one (needed to effectively control the disease) as more individuals are being educated over time.

We also assessed the impact of public health education on the outbreak. Our simulation shows that the possibility of curtailing the spread of the virus (bringing ℛ_*c*_ *<* 1) in the US is dependent on a very high education rate with high efficacy. The results obtained further showed the prospect of effective public health education programs in reducing both the cumulative and daily mortality of the novel coronavirus in the US. In particular, a 10% increase in education rate from the baseline value reduces the peak mortality by 66.3% by April 20, 2020, 26.7% by July 31, 2020, and 16% by December 8, 2020, when compared to the baseline scenario. However, a 40% increase in education rate from the baseline value reduces the peak daily mortality by 96.8% by April 7, 2020, 44.5% by July 24, 2020, and 54.8% by December 8, 2020. This result is consistent with what was obtained in [3, 5, 8, 18], where the universal use of face masks greatly curtailed community transmission of COVID-19 and brought the pandemic under very effective control.

The Centers for Disease Control and Prevention (CDC) at the early stage of the pandemic recommended the use of a face mask, social-distancing in public, and proper/frequent hand washing to curtail the spread of the novel coronavirus caused by SARS-CoV-2 [3, 6, 28]. Many state governments issued executive order mandating a face mask in public and restricting large gatherings of people. However, using a face mask and social-distancing in public places appears to be politicized in the US [29]. In particular, states like Georgia and Iowa barred Mayors and City Councils from introducing mask mandates, even as cases continues to rise in various counties in the state [30]. While many people strictly adhere to public health measures, others passionately ignore them. We ran simulations to show the impact of “loss of willingness” (fatigue rate) on both the cumulative and peak daily mortality. The result indicates that non-compliance to public health measures would positively impact the outcome of the pandemic. In particular, a 10% increase in fatigue rate from the baseline value increases the peak daily mortality by 6.9% by May 1, 2020, 10.1% by August 3, 2020, and 58.5% by December 8, 2020, when compared to the baseline scenario. However, a 40% increase in fatigue rate from the baseline value increases the peak daily mortality by 31.9% by April 20, 2020, 67% by July 27, 2020, and 254% by December 8, 2020, when compared to the baseline scenario. This result further supports the fact that states with less adherence to public health measures may experience more coronavirus cases and daily mortality than places where there is strict adherence [3, 5, 8].

## Data Availability

All data are publicly available through the John Hopkins University COVID-19 dashboard

https://github.com/CSSEGISandData/COVID-19

## Authors contributions

EAI, AR, RR, DI, JC, JH, MM, RP, ZD conceived the study; EAI designed the model; EAI collected and analysed the data; EAI and BO performed the numerical simulations; EAI, AR, RR, DI, JC, JH, MM, RP, ZD, FBA, BO, LA drafted the manuscript; EAI, AR, RR, DI, JC, JH, MM, RP, ZD, FBA, BO, LA revised the manuscript. All authors read and approved the final manuscript. Ethics approval was not required as all data used in the manuscript are publicly available.

## Appendix A Tables A Tables of variable descriptions, parameter descriptions, and parameter values

## Appendix B Reproduction Number

The model (2.1) has a disease-free equilibria (DFE) located within the hyperplane of disease free equilibria, given by

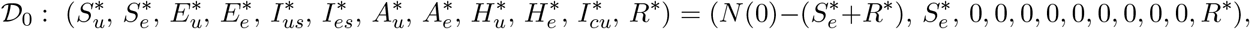

where *N* (0) is the initial total population size, 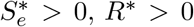, and 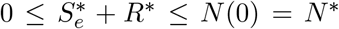. The next generation operator method [38, 39] can be used to analyse the asymptotic stability property of the disease-free equilibria, D_0_. In particular, using the notation in [38], it follows that the associated next generation matrices, *F* and *V*, for the new infection terms and the transition terms, are given, respectively, by

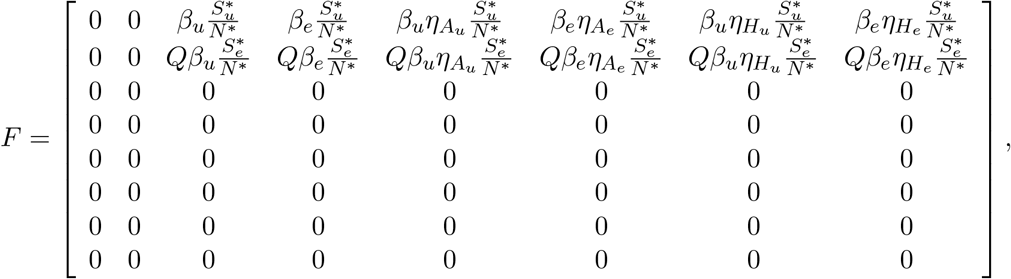

and,

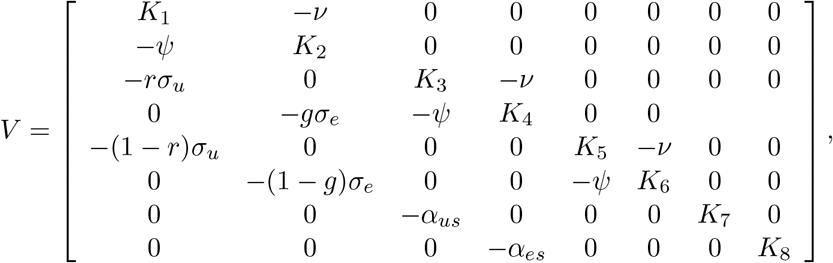

where *Q* = 1 − *ω, K*_1_ = *σ*_*u*_ + *ψ, K*_2_ = *σ*_*e*_ + *ν, K*_3_ = *α*_*us*_ + *γ*_*us*_ + *ψ* + *δ*_*us*_, and *K*_4_ = *α*_*es*_ + *γ*_*es*_ + *ν* + *δ*_*es*_, *K*_5_ = *γ*_*ua*_ + *ψ, K*_6_ = *γ*_*ea*_ + *ν, K*_7_ = *γ*_*hu*_ + *ϕ*_*hu*_ + *δ*_*hu*_, *K*_8_ = *γ*_*he*_ + *ϕ*_*he*_ + *δ*_*he*_. The control reproduction number is given by

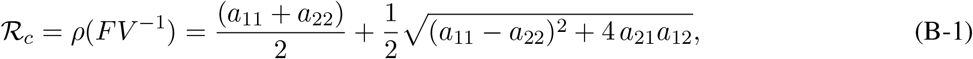

where,

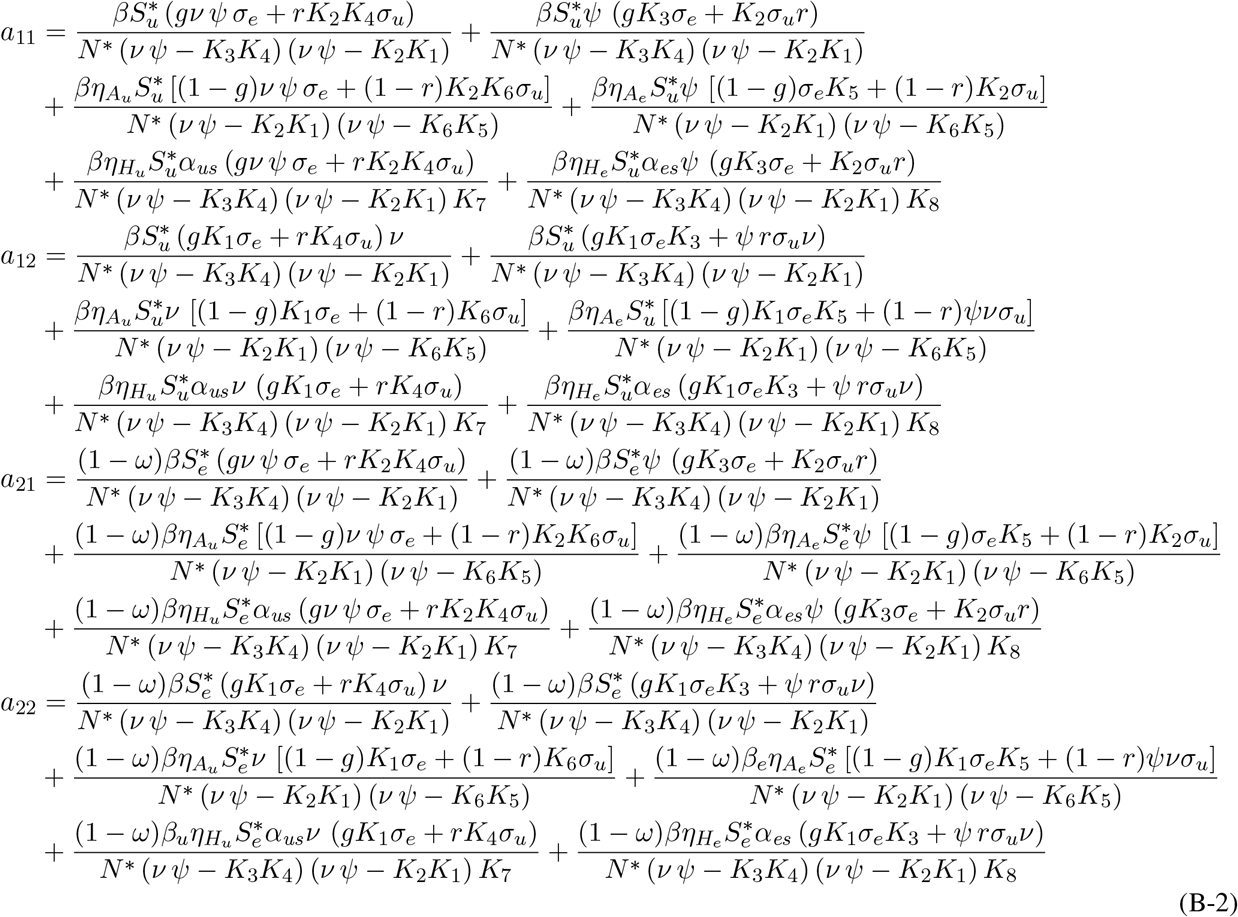

